# The public health impact of Paxlovid COVID-19 treatment in the United States

**DOI:** 10.1101/2023.06.16.23288870

**Authors:** Yuan Bai, Zhanwei Du, Lin Wang, Eric H. Y. Lau, Isaac Chun-Hai Fung, Petter Holme, Benjamin J. Cowling, Alison P. Galvani, Robert M. Krug, Lauren Ancel Meyers

**Affiliations:** WHO Collaborating Centre for Infectious Disease Epidemiology and Control, School of Public Health, LKS Faculty of Medicine, The University of Hong Kong, Hong Kong Special Administrative Region, China; Laboratory of Data Discovery for Health, Hong Kong Science and Technology Park, Hong Kong Special Administrative Region, China; Department of Genetics, University of Cambridge, Cambridge, UK; Department of Biostatistics, Epidemiology and Environmental Health Sciences, Jiann-Ping Hsu College of Public Health, Georgia Southern University, Statesboro, GA 30460, USA; Department of Computer Science, Aalto University, Espoo, FI 00076, Finland; Center for Computational Social Science, Kobe University, Nada, Kobe 657-8501, Japan; Center for Infectious Disease Modeling and Analysis, Yale School of Public Health, New Haven, CT, USA; Department of Molecular Biosciences, John Ring LaMontagne Center for Infectious Disease, Institute for Cellular and Molecular Biology, University of Texas at Austin, Austin, TX 78712, USA; Department of Integrative Biology, University of Texas at Austin, Austin, TX 78712, USA; Santa Fe Institute, Santa Fe, NM 87507, USA

**Author notes:** **Corresponding authors:** Lauren Ancel Meyers. Authors contributed equally (Yuan Bai, Zhanwei Du, Lin Wang, Eric H. Y. Lau).

**Keywords:** COVID-19, SARS-CoV-2 transmission, Paxlovid, public health impact, treatment, mathematical model

## Abstract

The antiviral drug Paxlovid has been shown to rapidly reduce viral load. Coupled with vaccination, timely administration of safe and effective antivirals could provide a path towards managing COVID-19 without restrictive non-pharmaceutical measures. Here, we estimate the population-level impacts of expanding treatment with Paxlovid in the US using a multi-scale mathematical model of SARS-CoV-2 transmission that incorporates the within-host viral load dynamics of the Omicron variant. We find that, under a low transmission scenario (*R*_*e*_ ∼ 1.2) treating 20% of symptomatic cases would be life and cost saving, leading to an estimated 0.26 (95% CrI: 0.03, 0.59) million hospitalizations averted, 30.61 (95% CrI: 1.69, 71.15) thousand deaths averted, and US$52.16 (95% CrI: 2.62, 122.63) billion reduction in health- and treatment-related costs. Rapid and broad use of the antiviral Paxlovid could substantially reduce COVID-19 morbidity and mortality, while averting socioeconomic hardship.

**Article Summary Line:** Mass treatment of symptomatic COVID-19 cases with antivirals that rapidly arrest SARS-CoV-2 replication would substantially reduce the spread and burden of the pandemic.

## Introduction

Antiviral drugs can substantially reduce morbidity and mortality from human infections. For example, antiretroviral therapy has prevented millions of HIV/AIDS deaths globally since the late 1980s (1). During the 2009 H1N1 influenza pandemic, oseltamivir was widely administered in the US (28.4 prescriptions/1,000 persons) (2); rapid treatment following symptom onset reduced the risk of hospitalization by an estimated 63% (95% CI: 17%–81%) (3). The influenza antiviral Baloxavir (Xofluza), which received approval for adults and children over 12 years of age by the Food and Drug Administration (FDA) in 2018 (4), blocks virus replication more rapidly and completely than oseltamivir (5). The reduction in viral load may reduce the risks of onward transmission while accelerating patient recovery. A counterfactual analysis suggests that treating even 10% of infected patients with baloxavir shortly after the onset of their symptoms would have prevented millions of infections and thousands of deaths in the US during the severe 2017-2018 influenza season (6). A fast-acting SARS-CoV-2 antiviral could similarly be deployed to curtail transmission on a population scale while directly saving lives (7).

Early efforts to develop SARS-CoV-2 antivirals focused on repurposing approved drugs for other pathogens that could be deployed without additional clinical trials. As of August 25, 2022, eleven SARS-CoV-2 therapies have been approved for emergency use in the US by the FDA (8,9). Remdesivir was the first repurposed drug to be fully approved in October of 2020. Originally developed to treat Ebola and Hepatitis C, it directly inhibits the SARS-CoV-2 RNA-dependent RNA polymerase (9,10). By December 2021, over 9 million patients were treated with Remdesivir worldwide, including 6.5 million patients in 127 middle- and low-income countries through Gilead’s voluntary licensing program (11). Sotrovimab was the second treatment to receive FDA emergency use authorization on May 26, 2021 (12). This monoclonal antibody therapy targets the spike protein and reduces the risk of hospitalization or death by an estimated 79% (13). On December 23, 2021, a newly developed drug, molnupiravir, received FDA emergency use authorization (14) and is estimated to reduce the risk of hospitalization or death by 30% (15). Paxlovid, which received FDA emergency authorization on December 22, 2021 for individuals over age 12, combines two different antiviral agents. The first component is a new drug, nirmatrelvir, which targets the SARS-CoV-2 3-chymotrypsin–like cysteine protease enzyme and inhibits viral replication. The second component is a repurposed drug, ritonavir, which was originally developed to treat HIV. Ritonavir inhibits CYP3A4 to slow the breakdown of nirmatrelvir in the patient (16). Treatment of symptomatic COVID-19 of patients with Paxlovid reduces hospitalization risks by an estimated 0.59 (95% CI: 0.48, 0.71) for adults aged 18–49 years, 0.40 (95% CI: 0.34, 0.48) for adults aged 50–64 years, and 0.53 (95% CI: 0.48, 0.58) for adults over 64 years(17). Paxlovid has proven effective against the Omicron variant (18). In January of 2022, the US ordered 20 million courses of Paxlovid to be delivered within nine months (19). The United Nations Children’s Fund (UNICEF) has ordered four million courses for distribution to lower income nations. Paxlovid is also licensed through the Medicines Patent Pool, allowing other manufacturers to produce low-cost generics. As of March 22, 2022, 35 companies in 12 nations across Latin America, the Middle East as well as South and East Asia have signed agreements to produce either the raw ingredients or the finished drug (20).

In this study, we analyze the population-level benefits of expanding the clinical use of Paxlovid to treat COVID-19 disease. By fitting a within-host model of viral replication to viral titer data from over 2000 SARS-CoV-2 patients, we provide early estimates for the efficacy of Paxlovid in curtailing viral load, depending on the timing of treatment after infection. Then, using a population-level SARS-CoV-2 transmission model, we estimate the impact of Paxlovid-based interventions on reducing the healthcare and economic burden of future COVID-19 epidemics. Specifically, we estimate the number of cases, hospitalizations and deaths, as well as healthcare costs averted under a range of transmission scenarios, in which we vary both the between-individual transmission rate of the virus and the proportion of cases who receive rapid treatment with Paxlovid. This two-level analytic framework can broadly support the rapid evaluation of antiviral-based mitigation strategies against COVID-19 and other respiratory viruses (6).

## Materials and Methods

### Within-host model of SARS-CoV-2 replication dynamics

We simulated SARS-CoV-2 virus kinetics in an infected individual and the effect of Paxlovid treatment using a standard target cell limited virus kinetic model (21,22). The deterministic model given by

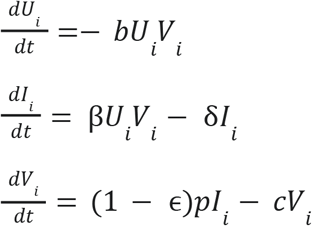

tracks the number of target cells at risk of infection (*U*_*i*_), infected cells (*I*_*i*_), and free viral particles (*V*_*i*_) (23) (**Fig. S1**). The rate at which free viral particles infect target cells is governed by the number of susceptible target cells, the number of free viral particles, and a fixed rate *b*. Viruses replicate at a rate *p* in infected cells; infected cells die at rate δ and free viral particles die at rate *c*. The model assumes that Paxlovid inhibits the replication of viruses within infected cells, with efficacy ε.

### Estimating the within-host model parameters

We fixed the initial number of viruses (*V*_0_) at 1/30 copy/mL (corresponding to a single viral particle per 30 mL of nasal wash in the upper respiratory tract (24)) and the initial number of target cells (*U*_0_) at 10^7^ (23). For treated patients, we assume that treatment is initiated eight days after infection, based on estimates for the Delta variant that the average time to symptom onset is five days (25) and average time from symptom onset to initiation of treatment is three days (16).

To estimate the five model parameters governing the viral load dynamics (i.e., the infection rate of susceptible cells [*b*], the rate at which infected cells die [δ], the rate at which active viruses were cleared [*c*], the virus production rate [*p*] and antiviral efficacy [ε]) we fitted the within-host model to the mean SARS-Cov-2 RNA titer (log10 copies/mL) at five time points (1, 3, 5, 10 and 14 days post initiation of treatment) measured across 1126 infected adults treated with a placebo during a clinical trial in late 2021 (16) and the mean SARS-Cov-2 RNA titer (log10 copies/mL) at five time points (1, 3, 5, 10 and 14 days post initiation of treatment) measured across 1120 infected adults who received Paxlovid in the same clinical trial (26). We set the initial viral load upon infection, *V*_0_, to correspond to 1 infectious virus particle in the upper respiratory tract (24) and assumed that the average viral load at the initiation of treatment is 10^6^ log10 copies/mL (16). We used the Stochastic Approximation Expectation-Maximization (SAEM) algorithm to estimate the five parameters (MONOLIX 2021R1) (27,28) and confirmed the convergence of estimates via trace plots.

### Modeling the infectiousness of treated and untreated cases

Following prior studies (29,30), we assumed that an individual’s infectiousness is logarithmically related to their viral titer, as given by:

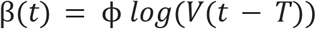

Where β(*t*) represents the infectiousness of the individual on day *t, V*(*x*) represents the viral load *x* days after becoming infected, *T* represents the day on which the individual was infected, and ϕ is a baseline transmission rate, which we estimated from epidemiological data, as described in (**Appendix, section S1**).

In the transmission model described below, we assume that the daily infectiousness of a case depends on whether or not they receive treatment and, if so, the time that treatment is initiated after symptom onset. To estimate the daily infectiousness of a given untreated or treated case, we first used the within-host model to simulate the viral load on each day of the infection, and set the viral load to zero when the estimated value dropped below the detection threshold of 100 (31). We then used the above equation to estimate the corresponding daily infectiousness.

### Modeling population-level SARS-CoV-2 transmission dynamics and the impacts of antiviral treatment

We developed a stochastic individual-based network model of SARS-CoV-2 transmission dynamics in which susceptible individuals can be infected by infected contacts (**Fig. S1**). The underlying contact network is described in (32); it includes 9961 individuals living in 5000 households with sociodemographic characteristics provided in the 2017 National Household Travel Survey (33) (**Appendix, sections S1 and S2**).

At every time point, each individual is in one of 11 possible states: unvaccinated susceptible (S_u_), vaccinated susceptible (S_v_), exposed (E), presymptomatic (P), symptomatic infectious prior to becoming eligible for Paxlovid treatment (Y), symptomatic treated (Y_T_), symptomatic untreated (Y_U_), asymptomatic infectious (A), recovered (R), hospitalized (H), or deceased (D). We assumed that hospitalized patients are isolated and not able to infect others. Upon infection, an susceptible individual progresses to the exposed state and then to either the presymptomatic or asymptomatic state, with probabilities τ and 1-τ, respectively. Asymptomatic cases recover without developing symptoms or seeking treatment. Presymptomatic cases progress to the symptomatic state at a rate ς, where they may be hospitalized according to published age-specific infection hospitalization rates (*h*_*a*_), and eventually recover or die from the infection, according to age-specific infection fatality rates (μ_*a*_). A fraction ρ of symptomatic cases receive Paxlovid, initiated an average of three days after symptom onset, which is assumed to reduce the risks of hospitalization (*φ*_*a*_) as well as the infectiousness of the individual. The infectiousness of a case depends on the timing of Paxlovid administration post infection, according to the daily infectiousness curves described in the preceding section. Vaccinated individuals initially have vaccine-derived immunity against infection ω_*B*_, symptomatic disease Ψ_*B*_, and death θ_*B*_ which wanes gradually post vaccination. Similarly, recovered individuals initially have infection-derived immunity against reinfection ω_*N*_, symptomatic disease Ψ_*N*_, and death θ_*N*_ which wanes more slowly than the vaccine-derived immunity. Individuals that are both vaccinated and recovered have the higher level of immunity of the two. Parameter definitions and values are provided in **Table 2, Appendix Table 1 and Appendix Table 2**. More details are in **Appendix S1**.

**Table 1.** Within-host parameter estimates. We fit the within-host model to the mean viral load dynamics reported from a clinical trial involving 2246 infected adults treated with either Paxlovid or a placebo (16) using nonlinear mixed-effects model method (27), a method that allows between-subject variability to improve the precision and accuracy of estimates (49). Values are means and 95% confidence intervals (CI) of parameter values across individuals.

| Parameter | Mean (95% CI) |
| --- | --- |
| Cell infection rate in $10^{-6}$ ml/Copies in days<sup>-1</sup> (b) | 3.92 (2.36, 6.5) |
| Infected cell death rate in days<sup>-1</sup> ($\delta$) | 0.63 (0.43, 0.92) |
| Virus production rate in Copies/ ml in days<sup>-1</sup> ($p$) | 3.28 (1.76, 5.9) |
| Virus death rate in days<sup>-1</sup> (c) | 2.21 (2.08, 2.35) |
| Antiviral efficacy ($\epsilon$) | 0.9938 (0.9895, 0.9964) |

**Table 2.** Between-host parameter estimates. We use the between-host model to project population-level impacts of Paxlovid treatment. Key parameter values used in the model are listed below, with more details in **Appendix Table 1**.

| Key parameter | Estimated Value |
| --- | --- |
| symptomatic proportion (%) ($\tau$) | 75 |
| Transition rate out of exposed state ($d^{-1}$) ($\sigma$) | 1/3 |
| Time lag between infection and recovery in days for asymptomatic patients ($d^{-1}$) ($\gamma_A$) | 1/9 |
| Time lag between symptom onset and recovery in days for symptomatic patients ($d^{-1}$) ($\gamma_T$) | 1/4 |
| transition rate from the pre-symptomatic to the symptomatic stage ($d^{-1}$) ($\varsigma$) | 1/2 |
| Age-specific efficacy of Paxlovid in reducing the hospitalization rate ($\varphi_a$) | [0.59 (95% CI: 0.48, 0.71), 0.59 (95% CI: 0.48, 0.71), 0.59 (95% CI: 0.48, 0.71), 0.40 (95% CI: 0.34, 0.48), 0.53 (95% CI: 0.48, 0.58)] for [0-4y, 5-17y, 18-49y, 50-64y, >65y] |
| Life expectancy (years) for age group $a$, adjusted assuming a 3% yearly discount rate ($\lambda_a$) | [30.3, 29.3, 25.8, 18.7, 12.9] for [0-4y, 5-17y, 18-49y, 50-64y, >65y] |

### Antiviral Treatment and Transmission Scenarios

We analyzed 24 different scenarios, each with an effective reproduction number (1.2, 1.5, 1.7, 2, 3 or 5) and Paxlovid treatment rate (20%, 50%, 80%, or 100%). For each scenario *s*, we compared four variations of the antiviral strategy: (i) no treatment (i.e., treatment rate set to zero), (ii) treatment with Paxlovid at the given treatment rate, (iii) treatment with a hypothetical antiviral that reduces infectiousness with the same efficacy as Paxlovid, but does not reduce severity, and (iv) treatment with a hypothetical antiviral that reduces severity with the same efficacy as Paxlovid, but does not reduce infectiousness. The last two variations allow us to separate the direct therapeutic benefits from the indirect transmission-blocking benefits of Paxlovid. To estimate the health and economic costs associated with each scenario, we ran 100 stochastic simulations of each of the four strategy variations and calculated the median and 95% prediction interval across simulations of the years of life lost (YLL) averted and monetary costs attributable to Paxlovid treatment.

### Estimating the Years of Life Lost (YLL) Averted and Monetary Costs

For each set of stochastic simulations, we estimated the years of life loss (YLL) averted for each antiviral strategy τ, by comparing it to the *no treatment* strategy, as follows:

1. Calculate the difference in incidence by age group as Δ_*a*,τ_ = *D*_*a*,0_ − *D*_*a*,τ_, where *D*_*a*,0_ and *D*_*a*,τ_ are total deaths in age group *a* produced by the no treatment and strategy τ simulations, respectively.
2. Estimate the YLL prevented by the strategy τ as 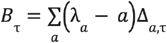 where λ _*a*_ denotes the future-discounted life expectancy for individuals of age *a*.

Similarly, we determined the incremental monetary costs for each strategy τ

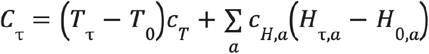

as given by where *T*_0_ and *T*_τ_ are the total number of treatment courses administered in the no treatment and strategy τ_*T*_ simulations, respectively, *c*_*T*_is the price of administering one course of antivirals, *H*_τ,*a*_ and *H*_0,*a*_ are the total number of hospitalizations in age group *a* in each simulation, and is the median COVID-19 hospitalization cost for age group *a*. The cost parameter values are given in **Table S3**.

The willingness to pay per YLL averted is the maximum price a society is willing to pay to prevent the loss of one year of life. Health economists have inferred from healthcare expenditure that the US is willing to pay US$100,000 per quality-adjusted life-year (34), of which YLL is one component. For a given willingness to pay for a YLL averted (θ), we calculated the net monetary benefit (NMB) of a strategy as

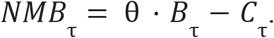

We determined the optimal strategy across a range of scenarios, each defined by the effective reproduction number (*R*_*e*_), willingness to pay, and cost of a vaccine.

### Sensitivity analyses

We assessed the robustness of the results with respect to two features of the model. We investigated three other functions relating viral load to infectiousness (i.e.., sigmoid, log-proportional, and step) (**Appendix Table 5**).

### Model validation

To validate our within-host viral replication mode, we compared model-estimated mean viral load trajectories for untreated and treated cases to corresponding clinical trial data for patients receiving placebo or Paxlovid treatment (1), and found that the observed mean decreases in viral load fall within the estimated 95% confidence intervals, and vice versa (Fig. 1).

**Fig. 1.**
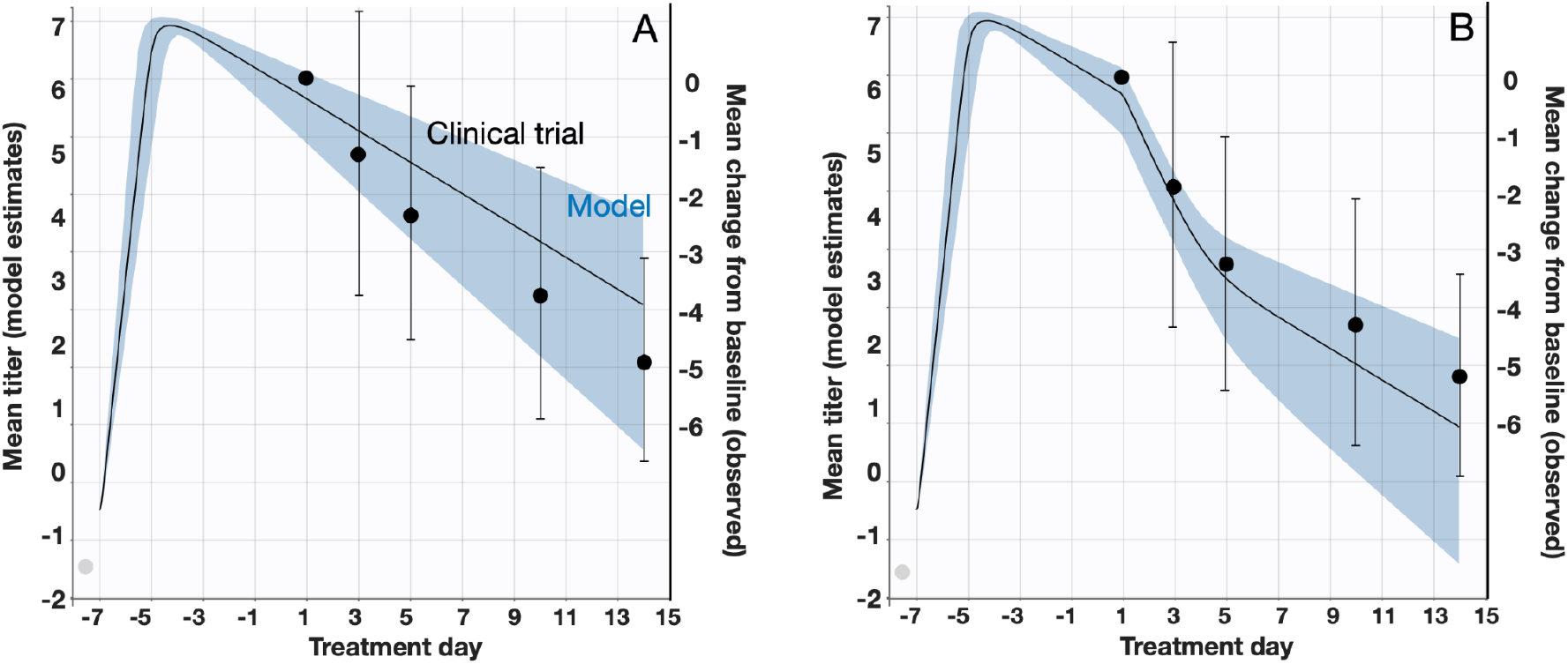
Estimated and observed viral load following treatment with (A) placebo or (B) Paxlovid. The left y-axes, black lines, and blue shading indicate the means and 95% confidence intervals of SARS-CoV-2 viral load (RNA log10 copies/mL) as estimated by the fitted within-host model. The right y-axes, black dots and error bars indicate the means and 95% confidence intervals of the decrease in viral load since the initiation of treatment as reported in a clinical trial in which 1126 COVID-19 patients received a placebo and 1120 patients received Paxlovid between July 16 and December 9, 2021 (16). Day one corresponds to the initiation of treatment. The gray circles denote the assumed initial viral load upon infection (V_0_) corresponding to one infectious virus particle in the upper respiratory tract (24).

To validate our transmission dynamic model, we compare model projections to observed incidence data during the early 2022 and late 2022 Omicron waves in the US (Fig. S3). For each of these waves, we fitted the model to reported case data to estimate the initial effective reproduction numbers and then simulated the expected reported infections, assuming a 25% case reporting rate (7).

## Results

By fitting the within-host model to the mean viral load dynamics reported from a clinical trial (**Fig. 1 and Table 1**), we estimate that the rate at which viral particles infect susceptible cells (β) is 3.92 (95% CrI: 2.36, 6.5) × 10^−6^ (copies/mL)^−1^ day^−1^, the clearance rate for infected cells (δ) is 0.63 (95% CrI: 0.43, 0.92) day^−1^, the rate at which infected cells release virus (p) is 3.28 (95% CrI: 1.76, 5.9) copies/mL day^-1^cell^-1^, and the rate at which free virus particles are cleared (c) is 2.21 (95% CrI: 2.08, 2.35) day^−1^. Treatment with Paxlovid is estimated to repress viral replication by 99.38% (95% CrI: 98.95%, 99.64%) per day.

We estimated the number of cases, hospitalizations and deaths, as well as healthcare costs, averted under a range of transmission scenarios, in which we vary both the between-individual transmission rate of the virus and the proportion of cases who receive rapid treatment with Paxlovid (**Fig. 2 and Fig. 3**). Under a low transmission scenario in which the effective reproduction number (*R*_e_) of the virus is 1.2, we estimate that treating 20% of symptomatic cases with Paxlovid would avert 9.85 (95% CrI: 3.03, 21.12) million cases, 0.26 (95% CrI: 0.03, 0.59) million hospitalizations, 30.61 (95% CrI: 1.69, 71.15) thousand deaths in the US over a 300-day period (**Appendix Table 4**). Assuming a cost of US$530 per course of treatment (35) and willingness to pay per year of life lost (YLL) averted of US$100,000, we estimate that the optimal strategy is always the highest treatment rate achievable. A 20% treatment rate would be expected to yield a net monetary benefit (NMB) of US$52.16 (95% CrI: 2.62, 122.63) billion averted. To separate the direct (therapeutic) benefits of Paxlovid treatment from its indirect (transmission-reducing) impacts, we conducted two additional analyses, one assuming the drug reduces severity but not infectivity and another assuming the opposite (**Appendix Table 4**). Assuming an *R*_e_ of 1.2, we estimate that direct therapeutic effects of treating 20% of symptomatic cases with Paxlovid would not impact the overall attack rate but would avert 0.10 (95% CrI: -0.13, 0.40) million hospitalizations and 14.39 (95% CrI: -19.47, 48.11) thousand deaths over a 300 day period, resulting in a NMB of US$24.10 (95% CrI: -34.98, 84.22) billion. The reduced infectivity of the treated cases would be expected avert an additional 9.88 (95% CrI: 3.03, 21.19) million infections, 0.13 (95% CrI: -0.13, 0.53) million hospitalizations, and 18.68 (95% CrI: -14.14, 58.52) thousand deaths, resulting in a NMB of US$26.57 (95% CrI: -32.77, 103.74) billion.

**Fig. 2.**
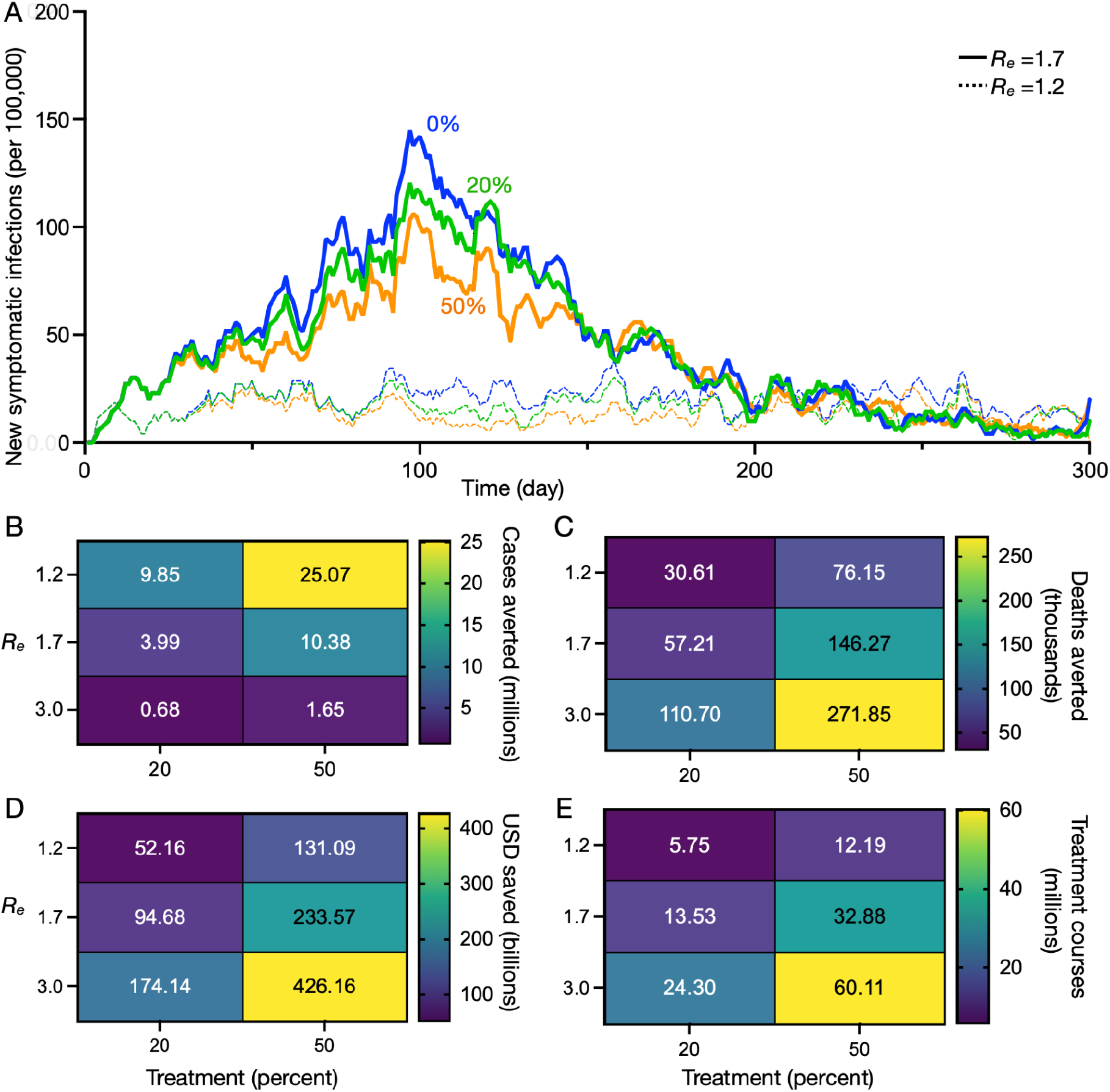
Projected health and economic impacts of a large-scale SARS-CoV-2 antiviral campaign over 300 days in the US, across a range of transmission and treatment scenarios. (A) Estimated incidence of symptomatic SARS-CoV-2 infections across three treating scenarios: 0%, 20%, or 50% of symptomatic cases receive a five-day Paxlovid regimen started within three days of symptom onset (indicated by line color). Dashed and solid curves correspond to effective reproduction numbers of 1.2 and 1.7, respectively. The four heat maps provide estimates across three different effective reproduction numbers (rows) and two different treatment rates (columns), of the following quantities: (B) number of cases averted (millions), (C) number of deaths averted (thousands), (D) net monetary benefit (NMB) in billions of USD assuming a treatment course cost of US$530 and WTP per YLL averted of US$100,000, and (E) number of courses of Paxlovid administered (millions). Cell color and value indicates the median estimate across 100 pairs of stochastic simulations (treatment vs no treatment simulations). The results are all scaled assuming a US population of 328.2 million people (50).

**Fig. 3.**
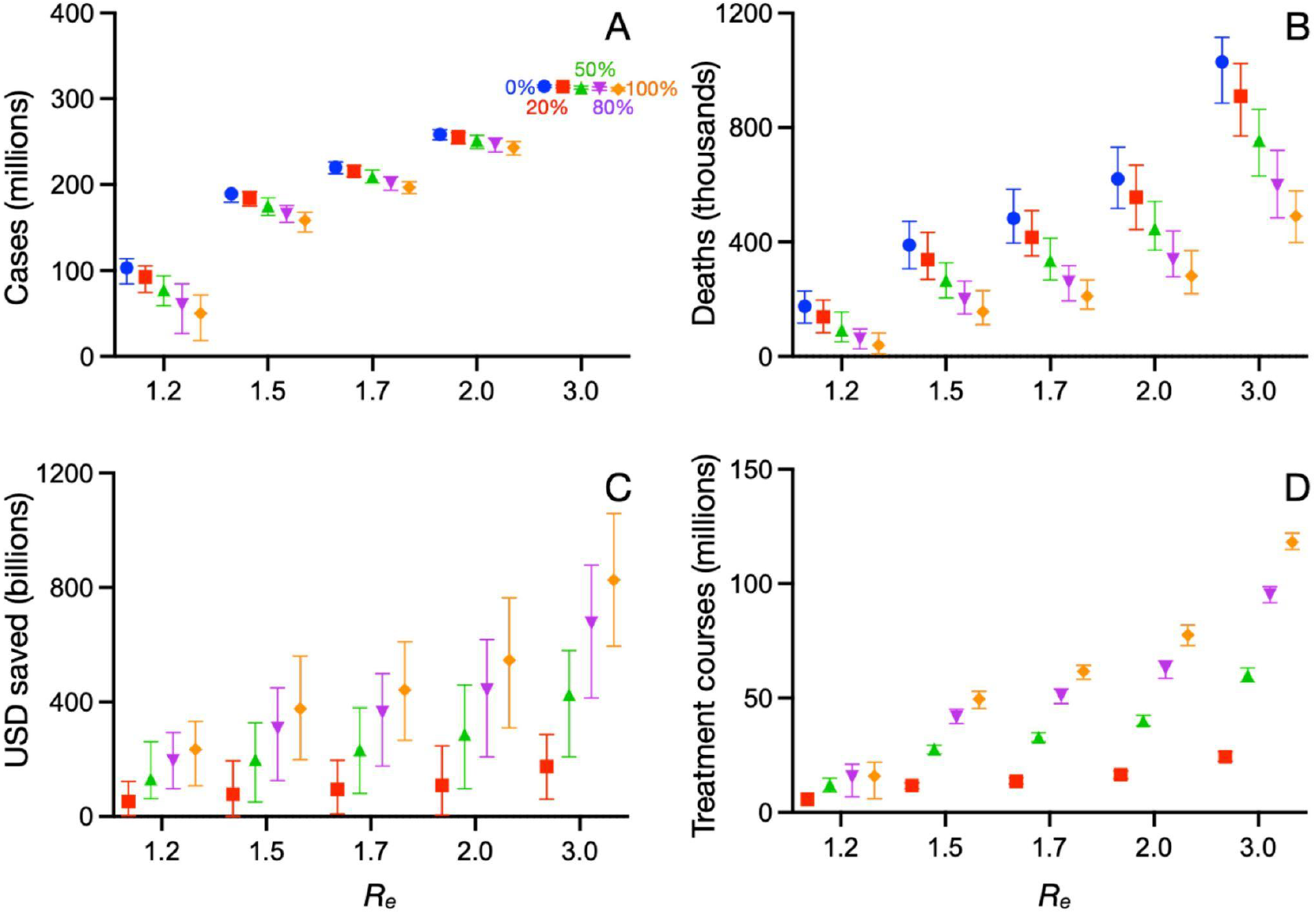
Projected health and economic impacts of a large-scale SARS-CoV-2 antiviral campaign over 300 days in the US, across a range of transmission and treatment scenarios. For five different effective reproduction numbers (1.2, 1.5, 1.7, 2, 3) and four different treatment scenarios (0%, 20%, 50%, 80%, or 100% of symptomatic cases start a five-day course of Paxlovid within three days of symptom onset), we estimate the median and 95% credible intervals in (A) number of cases infected (millions), (B) number of deaths (thousands), (C) net monetary benefit (NMB) in billions of USD assuming a treatment course cost of US$530 and willingness to pay (WTP) per year of life lost (YLL) averted of US$100,000, and (D) number of courses of Paxlovid administered (millions). Distributions are based on 100 stochastic simulations for each scenario. The results are all scaled assuming a US population of 328.2 million people (50).

## Discussion

The widespread administration of Paxlovid would not only improve outcomes in treated patients but would concomitantly reduce risks of onward transmission. In this population-level assessment of expanding rapid treatment of symptomatic COVID-19 infections with Paxlovid, we find that the direct (therapeutic) effects of treatment would significantly reduce both mortality and socioeconomic costs. Importantly, the indirect (transmission-blocking) effects would be expected to reduce burden by just as much, as well as substantially reduce the overall attack rate **(Appendix Table 4)**. We would expect mass treatment campaigns to have even greater health and economic impacts in countries that have adopted zero-COVID strategies and thus have lower levels of population-level immunity than the US (36).

Drugs like Paxlovid could profoundly reduce the severity of COVID-19 and facilitate the global transition to manageable coexistence with the virus. However, providing equitable and effective global access to SARS-CoV-2 antivirals would require both ample supplies and broad-reaching test and treat programs. The pharmaceutical industry and global health agencies are racing to produce sufficient quantities of Paxlovid to treat a large fraction of symptomatic cases (19). Online healthcare services (e.g., telemedicine) and community test-to-treat programs (37), such as those piloted in Pennsylvania and New Jersey (38), could be expanded nationally, and even globally, to accelerate and broaden access to antivirals (39). For example, in 2020, China began an initiative to expand remote internet-based COVID-19 care (40). They established 1500 internet hospitals (either extending existing hospitals or new institutions) between 2019 to 2021 (41). The new services included follow-up consultations for common ailments (42) and served over 239 million patients between December 2020 and June 2021 (43). In addition, avoiding in-person testing and administration of infected individuals reduces risks of SARS-CoV-2 transmission by patients to healthcare providers.

We highlight three limitations of our analyses that could be addressed as additional epidemiological and clinical trial data become available. First, our fitted within-host model slightly overestimated viral levels for patients treated with placebo and underestimates those for patients receiving Paxlovid. The discrepancies may stem from limitations in the model structure or from unmodeled variation in viral kinetics and treatment efficacy across age or risk groups. In estimating model parameters, we considered only the mean in viral load of patients from 20 countries (6,44) (**Fig. 1**). Incorporating such variability would allow us to analyze age- or risk-prioritized interventions and improve our estimates of the health and economic benefits of mass treatment. Second, we did not consider the emergence and spread of Paxlovid-resistant viruses, which could significantly undermine the utility of new drugs and exacerbate epidemics on a population level (45). Conversely, suppressed viral replication attributable to Paxlovid may limit viral evolution in treated patients. Depending on the immunological conditions of the individual and population, reducing opportunities for viral growth and mutations may hinder the emergence of new variants (46). Third, we did not incorporate a number of economic, social, and logistical factors that may impact the expansion of Paxlovid treatment, including commercial impediments faced by the pharmaceutical companies that manufacture the drug (47), the costs of administering tests prior to treatment, and low levels of uptake stemming from misinformation, limited healthcare access, or pandemic fatigue. For example, in the 2009 H1N1 pandemic, only 40% of cases sought medical care within 3 days post symptom onset (48). In conclusion, fast-acting antiviral drugs like Paxlovid can serve as invaluable tools to mitigate COVID-19 epidemics. By increasing supplies and the infrastructure needed for rapid and equitable distribution, such drugs could substantially mitigate the health and societal burdens of COVID-19.

## Data Availability

All data produced in the present study are available upon reasonable request to the authors

## Acknowledgements

Financial support was provided by the AIR@InnoHK Programme from Innovation and Technology Commission of the Government of the Hong Kong Special Administrative Region, the US National Institutes of Health (grant no. R01 AI151176), the Centers for Disease Control and Prevention COVID Supplement (grant no. U01IP001136), and Health and Medical Research Fund, Food and Health Bureau, Government of the Hong Kong Special Administrative Region (grant no. 21200632).

## Ethics committee approval

Not applicable.

## Contributors

Y.B., Z.D., B.J.C., A.P.G., R.M.K, and L.A.M., designed research; Y.B., and Z.D. performed research; L.W., E.H.Y.L., I.C.H.F., and P.H. contributed analytic method comments. Y.B., Z.D., L.W., E.H.Y.L., I.C.H.F., P.H., B.J.C., A.P.G., R.M.K, and L.A.M. wrote the paper.

## Data sharing

The computer code is available in Github https://github.com/ZhanweiDU/Pax.

## Author Bio

Dr. Zhanwei Du is a research assistant professor in the School of Public Health, LKS Faculty of Medicine, The University of Hong Kong, Hong Kong Special Administrative Region, China. He develops mathematical models to elucidate the transmission dynamics, surveillance, and control of infectious diseases.

