## Appendix for "The public health impact of Paxlovid COVID-19 treatment in the United States"

[†] Authors contributed equally

### Appendix

#### S1. Between-host model of SARS-CoV-2 transmission and treatment with Paxlovid-like drug

The infectiousness of a case depends on the timing post infection and the type of contact (i.e., household or non-household). To parameterize the baseline infectiousness of symptomatic cases, we first calibrated the transmission rate $\beta_h$ between individuals of the same household to produce a household secondary attack rate of 35%, according to recent estimates in the US (1). To model the changing infectivity of individual cases throughout their infection, $\beta(t)$, we assumed the viral load of an infected individual decreases to zero before the 50-th day post infection, the overall transmission of $\beta_h$ over days is $\xi_h \sum_{t=0}^{50} \beta(t)$, where the coefficient $\xi_h = \frac{35\%}{\sum_{t=0}^{50} \beta(t)}$.

We initialized each stochastic simulation assuming vaccination levels reported in January of 2022 (**Tables S1**). In each simulated epidemic without treatment, we estimated the effective reproduction number ($R_e$) by calculating the average number of secondary infections across the selected 1% randomly infected individuals during the first 100 days of the simulation. To parametrize the model to produce a specific value of $R_e$, we used an interior point algorithm to find the coefficient $\xi_{nh}$ of a non-household (e.g., schools, workplaces, and other venues) transmission rate ($\beta_{nh}(t) = \xi_{nh}\beta(t)$) that minimizes the mean square error between the target $R_e$ and the $R_e$ produced by 100 simulated epidemics. We denoted the effective reproduction number for scenarios with vaccination by $\overline{R_e}$. Simulations initiate both an antiviral rollout and a *status quo* strategy in which no cases are treated.

In the start of the simulation, we assumed proportions of population vaccinated or recovered according to estimates from US Centers for Disease Control and Prevention

(Appendix Table 1), and 1% population in exposed state. To estimate the number of previously vaccinated individuals and the rate of their most recent dose, we simulated vaccination rates based on reported uptake in the US from 2020 to 2022 (2). For each previously vaccinated individual, we randomly selected the date of their first dose ($t_1$) based on the reported age-specific vaccine administration rates, starting on October 29, 2021 (3) for children between 5 and 11 years old, May 10, 2021 for children between 12 and 15 years old (4), and December 13, 2020 for all others. We then randomly determined whether and when an individual receives their second primary dose and first booster based on CDC-recommended waiting periods and reported rates of uptake. Specifically, we assumed second doses are administered beginning three weeks after the first dose and the window for boosters depends on the timing of the booster dose, with a minimum gap of eight months for individuals receiving their booster dose before September 23, 2021 (5), six months between September 24, 2021 and January 3, 2022 (6), and five months after January 4, 2022 (7). We initialized immunity in our simulations using the dates of the last dose received for each vaccinated individual (**Appendix Table 1**) (8).

For the previously infected individuals, we estimated their times of recovery. Specifically, we collected the daily population proportion of confirmed cases in the USA from 2021 to 2022 from Our World In Data (2). For each individual infected previously at the start of the simulation, we estimated the date of the previous infection ($t_{infect}$) by taking draws from the distribution of the daily population proportion of cases between January 29, 2021 to January 29, 2022. We considered the time of recovery as ($t_{infect}$ + 9), where 9 days is the average time lag between infection and recovery (9).

At the start of each simulation, we assumed that portions of the population have been previously vaccinated or infected, according to estimates from US Centers for Disease Control and Prevention (**Appendix Table 1**). We initially moved one percent of the fully susceptible/vaccinated population (i.e., not previously infected) into the newly infected (exposed) compartment, which corresponds to approximately 0.6% of the total population. In each simulation, the effective reproduction number ($R_e$) corresponds to the average number of secondary infections at the outset of the epidemic. When calibrating the transmission rate to achieve a specified $R_e$, we tracked the number of secondary infections caused by a randomly selected 1% of infected and untreated individuals during the first 100 days of the simulation (**SI Appendix, section S1**). We assumed age-stratified estimates for Paxlovid's efficacy at preventing hospitalizations (10) (**Appendix Table 1**) and incorporated uncertainty by sampling efficacies for each simulation from triangular distributions with mean, lower bound, and upper bound equal to the estimated mean, 95% CI lower bound, and 95% CI upper bound, respectively. To estimate therapeutic benefits of the drug via pairs of simulations, we enforced the same sequence of random numbers in each simulation.

### S2. Individual-based network construction

The individual-based SARS-CoV-2 infection dynamic model assumes that the virus spreads through a fixed contact network consisting of 9961 individuals and 124,878 contacts between those individuals. We populated our network by first constructing 5000 households. The size and age composition of each household is based on a randomly sampled household from among the 129,697 households included in 2017 National Household Travel Survey (11). We assumed that households are fully connected (i.e., all nodes in the same household are linked by edges). We constructed random links between individuals in different households based on reported age-specific contact rates in the US, stratified into age bins of 5–17, 18–49, 50–64, and over 65 years (12). Specifically, to determine the number of contacts a node in age group $a_i$ has with nodes in age group $a_j$ we draw random deviates from Poisson distributions centered at the mean number of contacts between $a_i$ and $a_j$. The resulting network includes 5000 households, 2019 nodes (people), and degrees (numbers of edges per node) that roughly follow a gamma distribution with shape and scale parameters of 3.69 and 3.41, respectively. We directly scaled our results to the 2019 US population of 328 million (13).

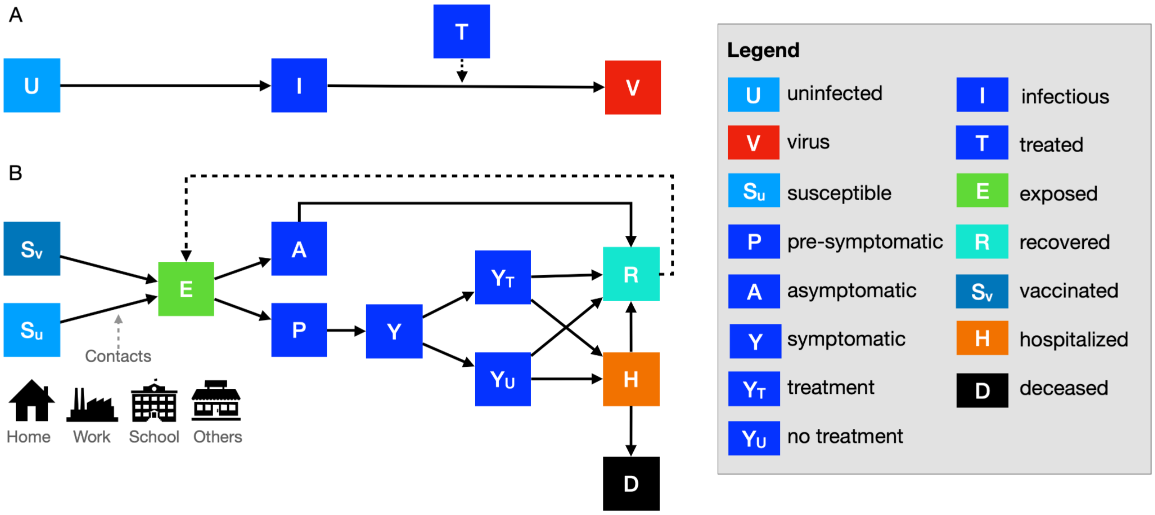


**Fig. S1. Diagrams of the within-host and between-host models.** (A) To estimate changes in infectivity following treatment with Paxlovid (T), we used a model that tracks the changing number of uninfected cells (U), infectious cells (I) and free viral particles (V) in an infected case, with and without treatment. (B) We projected population-level impacts of Paxlovid treatment using a stochastic individual-based model of SARS-CoV-2 transmission that considers age-specific risks and contact patterns in households, schools, workplaces, and other venues. Upon infection, susceptible ($S_u$) and vaccinated ($S_v$) individuals progress to exposed (E), asymptomatic infectious (A) or presymptomatic (P) and then to either symptomatic infectious (Y) with ($Y_T$) or without ($Y_U$) Paxlovid treatment. A fraction of symptomatic cases with or without treatment will be recovered (R) or hospitalized (H), and a subset of those will die (D). All asymptomatic cases eventually progress to a recovered class (R) where they remain protected from future infection. As immunity wanes, recovered individuals return to exposed (E) by reinfection.

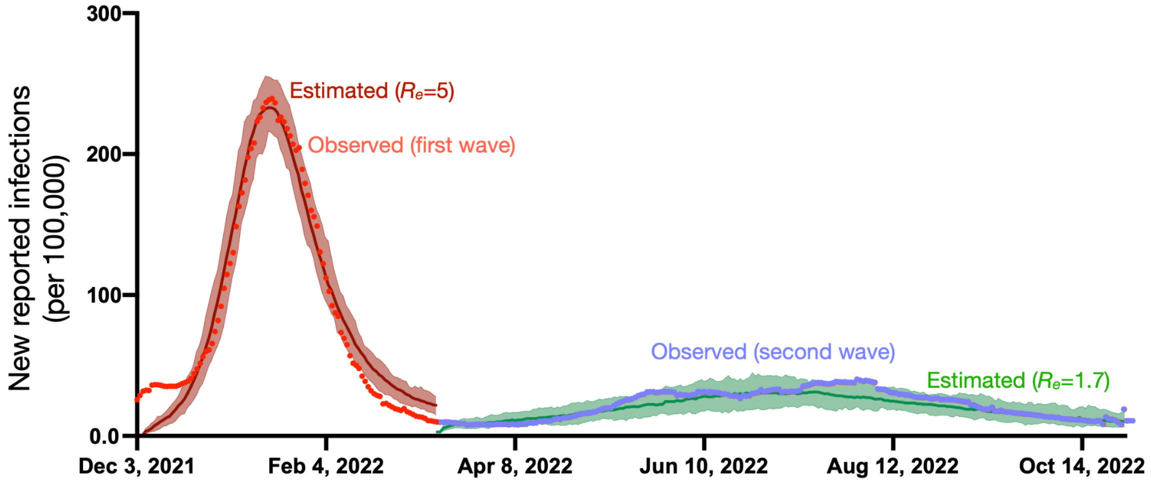


**Fig. S2. Comparison of observed** (8) **to estimated SARS-CoV-2 incidence during two Omicron waves in the US.** Model estimates assume an initial effective reproduction number of 5.0 for the first large Omicron wave in the US (December 3, 2021 to March 12, 2022) and a reproduction number of 1.7 for the second smaller wave (March 13, 2021 to October 31, 2022), and assume a case reporting rate of 25% [8]. Points indicate reported incidence data; lines and shading correspond to the median and 95% prediction interval across 100 stochastic simulations for each wave.

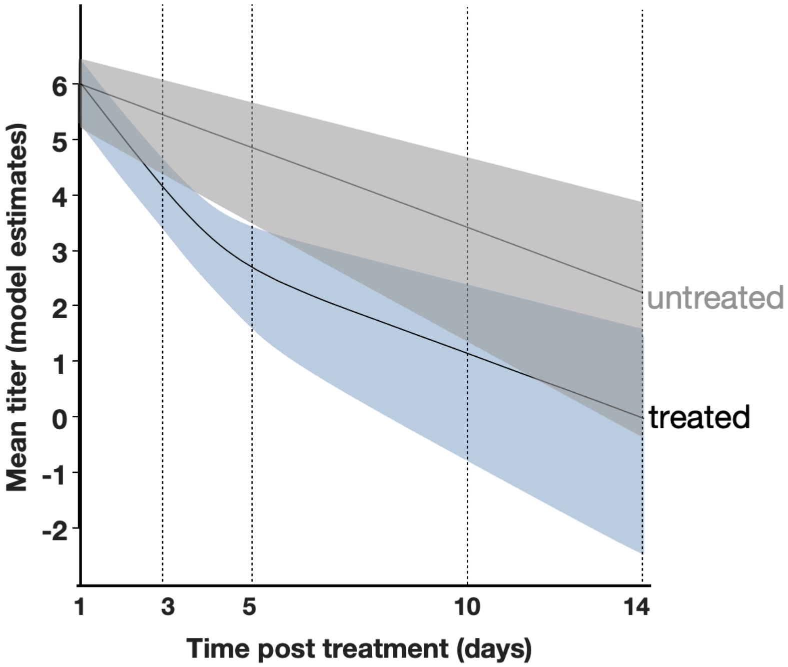


**Fig. S3. Comparison between the estimated mean viral loads for treated versus untreated cases.** Lines and shading indicate the means and 95% confidence intervals of SARS-CoV-2 viral load (RNA log10 copies/mL) as estimated by the fitted within-host model. Day one corresponds to the initiation of treatment.

**Appendix Table 1. Parameter values.**

| Parameters | Values | Source |
|---|---|---|
| *N*: number of individuals in the population | 9961 individuals in 5000 households | |
| Initial percent vaccinated as of January 29, 2022 for [0-4y, 5-17y, 18-49y, 50-64y, >65y] | At least one dose: [0, 42.4%, 77.2%, 89.6%, 97.4%]<br><br>One dose only: [0%, 4.50%, 2.30%, 1.70%, and 1.20%]<br><br>Primary series only (two doses): [0%, 37.90%, 29.60%, 25.80%, and 21.30%]<br><br>Primary and first booster: [0%, 0%, 45.30%, 62.10%, and 74.90%] | (8) |
| Initial number of infections (in exposed state) | 1% of the susceptible population | Assumed |
| Initial proportion of recovered | [39.34%, 60.81%, 50.61%, 48.38%, and 35.88%] for [0-4y, 5-17y, 18-49y, 50-64y, >65y] | (1) 77 million, 129 million, and 220 million total cases were reported in the US on 29 January 2021, 30 September 2021, and 29 January 2022 (2).<br>(2) US CDC estimates [35.49%, 54.86%, 45.66%, 43.65%, 32.36%] for [0-4y, 5-17y, 18-49y, 50-64y, >65y] |

| | | |
|---|---|---|
| | | from February 2020 to September 2021 (14).<br>(3) We estimated the proportion of infected cases in the past year (between January 29, 2021 and January 29, 2022) used at the start of simulations as (220-77)/129.*[35.49%, 54.86%, 45.66%, 43.65%, 32.36%] = [39.34%, 60.81%, 50.61%, 48.38%, 35.88%], for the four age groups, respectively. |
| τ: symptomatic proportion (%) | 75 | (15) |
| ρ: treatment proportion (%) | | Assumed |
| $\xi_{nh}$: baseline transmission rate | $\xi_{nh}$ is 0.0004, 0.0005, 0.0006, 0.0008, and 0.0013 for $R_e$ of 1.2, 1.5, 1.7, 2, and 3, respectively. | Calibrated to $R_e$. |
| σ: transition rate out of exposed state ($d^{-1}$) | 1/3 | (16) |
| $\gamma_A$: transition rate between infection and recovery in days for asymptomatic patients ($d^{-1}$) | 1/9 | (9) |
| $\gamma_Y$: transition rate between symptom onset and recovery in days for symptomatic patients ($d^{-1}$) | 1/4 | (17,18) |

| | | |
|---|---|---|
| ς: transition rate from the pre-symptomatic to the symptomatic stage ($d^{-1}$) | 1/2 | (16) |
| $h_a$: age-specific proportion of symptomatic cases that are hospitalized | [0%, 0.025%, 2.672%, 9.334%, 15.465%]<br>for [0-4y, 5-17y, 18-49y, 50-64y, >65y] | (19) |
| $\varphi_a$: age-specific efficacy of Paxlovid in reducing the hospitalization rate | [0.59 (95% CI: 0.48, 0.71), 0.59 (95% CI: 0.48, 0.71), 0.59 (95% CI: 0.48, 0.71), 0.40 (95% CI: 0.34, 0.48), 0.53 (95% CI: 0.48, 0.58)] for [0-4y, 5-17y, 18-49y, 50-64y, >65y] | (10)<br>The study provides estimates for adults over age 18y. We assumed that efficacy for children under 18 is the same as that for adults aged 18-49y. |
| $\eta$: transition rate from treatment to hospitalized ($d^{-1}$) | $1/(5.9-1/\gamma_T)$ | 5.9 days on average from symptomatic to hospitalized (20) |
| $\gamma_T$: transition rate from symptomatic to treatment ($d^{-1}$) | 1/3 | an average of 3 days between COVID-19 symptom onset and the initiation of Paxlovid treatment (21) |
| $\mu_a$: age-specific mortality rate for hospitalized cases | [0.48%, 0.48%, 5.68%, 10.82%, 16.15%] for [0-4y, 5-17y, 18-49y, 50-64y, >65y] | (22) |
| $\gamma_d$: transition rate from hospitalized to deceased for cases that succumb ($d^{-1}$) | 0.128 | (23) |

| | | |
|---|---|---|
| $\gamma_h$: transition rate from hospitalized to recovered for cases discharged alive ($d^{-1}$) | 0.091 | (23) |
| $\lambda_a$: life expectancy (years) for age group *a*, adjusted assuming a 3% yearly discount rate | [30.3, 29.3, 25.8, 18.7, 12.9] for [0-4y, 5-17y, 18-49y, 50-64y, >65y] | (24) |

**Appendix Table 2. Parameters governing waning of immunity following vaccination and infection with respect to the SARS-CoV-2 Omicron variant.** Vaccine-related parameter values are based on estimates for the BNT162b2 (Pfizer) vaccine.

| Time since most recent vaccination | Immunity following vaccination | | |
|---|---|---|---|
| | **Reduction in susceptibility to infection ($\omega_B$)** | **Reduction in likelihood of developing symptoms following infection ($\psi_B$)** | **Reduction in risk of mortality following infection ($\theta_B$)** |
| 1 week | 64%* | 66.9% (73) | 91.9% * |
| 2 to 4 weeks | 64%* | 67.2% (73) | 91.9%* |
| 5 to 9 weeks | 42.8%* | 55.0% (73) | 91.9%* |
| >= 10 weeks | 22%* | 45.7% (73) | 91.9%* |
| **Time since recovery** | **Immunity following infection** | | |
| | **Reduction in susceptibility to infection ($\omega_N$)** | **Reduction in likelihood of developing symptoms following infection ($\psi_N$)** | **Reduction in risk of mortality following infection ($\theta_N$)** |
| 1 week | 83.1%+ | 92.1%++ | 98.1%++ |
| 2 to 4 weeks | 83.1%+ | 92.1%++ | 98.1%++ |
| 5 to 9 weeks | 73.1%+ | 89.2%++ | 98.1%++ |
| >= 10 weeks | 63.3%+ | 87.0%++ | 98.1%++ |

* Since direct estimates of $\omega_B$ and $\theta_B$ for vaccine booster doses against the Omicron variant are not available, we extrapolated from Ref. (25) which estimates $\omega_B$ for boosters against Omicron and other studies that simultaneously estimate vaccine efficacy against infection, symptomatic infection, and mortality for the Pfizer-BioNTech BNT162b2 vaccine earlier in the pandemic. In Ref. (26), vaccine efficacy against infection is estimated to be 64% when efficacy against symptomatic disease reaches 67% (21 days after vaccination). In Ref. (27), vaccine efficacy against infection is estimated to be 42.8% when

efficacy against symptomatic disease reaches 52.4% (14 days after the initial vaccine dose). In Ref. (26), vaccine efficacy against infection is estimated to be 22% when efficacy against symptomatic disease reaches 44% (14 days after the initial vaccine dose). In Ref. (28), vaccine efficacy against mortality is estimated to be 91.9% when efficacy against symptomatic disease reaches 66.3% (20 days after the second vaccine dose).

** Since direct estimates for $\psi_N$ against the Omicron variant are not available, we extrapolated from Ref. (29) which estimates $\omega_N$ against Omicron and Ref. (30), which reports that vaccine efficacy against symptomatic disease is 97% when efficacy against infection reaches 79% (14 days after the second vaccine dose).

$^{+}$ Using an estimated adjusted hazard ratio of SARS-CoV-2 infection following natural infection versus BNT162b2 vaccination of 0.47 (95% CI: 0.45–0.48), we scaled $\omega_N$ based on $\omega_B$ (31).

$^{++}$ Using an estimated adjusted hazard ratio of severe SARS-CoV-2 infection following natural infection versus BNT162b2 vaccination of 0.24 (95% CI: 0.08–0.72), we scaled $\psi_N$ based on $\psi_B$ (31) and $\theta_N$ based on $\theta_B$..

**Appendix Table 3. Cost parameters.**

| **Parameter** | **Value (USD)** |
|---|---|
| Cost of administering a treatment course ($c_T$) by taking Paxlovid | \$530 per course (32) |
| Median COVID-19 hospitalization cost by age group ($c_{H,a}$) | [\$21,847, \$21,847, \$19,681, \$23,157, \$18,806] for [0-4y, 5-17y, 18-49y, 50-64y, >65y] (33) |

**Appendix Table 4. Projected cases, hospitalizations and deaths averted in the US, net monetary benefit (billion USD), and treatment courses administered in a large-scale SARS-CoV-2 antiviral campaign, across four treatment rates and five transmission scenarios (effective reproduction numbers from 1.2 to 5).** Assuming a treatment course cost of US$530 and willingness to pay (WTP) per year of life lost (*YLL*) averted of US$100,000, for each reproduction number and treatment rate, we estimated the median and 95% credible intervals based on 100 pairs of stochastic simulations (treatment vs. no treatment simulations). To separate the direct therapeutic benefits of the drug from the indirect transmission-blocking impacts of treatment, we analyzed an alternative model in which the drug improves patient outcomes but does not impact infectivity.

| **Outcome** | $R_e$ | **Treatment rate (% symptomatic cases)** | **Median (95% CrI)** | | |
|---|---|---|---|---|---|
| | | | **Base model** | **Alternative model: direct (therapeutic) effects only** | **Alternative model: indirect (transmission reducing) effect only** |
| No. of cases averted (million) | 1.2 | 20% | 9.88 (3.03, 21.19) | -0.07 (-3.82, 1.91) | 9.85 (3.03, 21.12) |
| | | 50% | 25.14 (12.88, 45.11) | -0.20 (-4.84, 2.37) | 25.07 (12.59, 41.19) |
| | | 80% | 42.44 (25.30, 80.72) | -0.35 (-5.34, 2.90) | 41.96 (25.24, 67.45) |
| | | 100% | 53.66 (36.77, 81.94) | -0.36 (-6.19, 3.26) | 53.10 (36.67, 81.88) |
| | 1.5 | 20% | 5.14 (0.07, 9.32) | -0.18 (-2.14, 0.99) | 5.07 (-0.03, 9.29) |
| | | 50% | 13.77 (7.55, 19.54) | -0.25 (-3.69, 1.98) | 13.77 (7.51, 19.21) |
| | | 80% | 23.85 (16.38, 30.21) | -0.51 (-4.68, 2.54) | 23.71 (16.21, 29.92) |
| | | 100% | 30.94 (23.43, 41.35) | -0.81 (-4.61, 2.17) | 30.59 (23.39, 40.69) |
| | 1.7 | 20% | 3.99 (0.07, 8.37) | -0.16 (-2.14, 1.45) | 3.99 (0.00, 8.30) |
| | | 50% | 10.58 (6.16, 16.70) | -0.33 (-2.64, 1.91) | 10.38 (5.77, 16.70) |
| | | 80% | 18.29 (10.54, 24.41) | -0.56 (-2.73, 1.52) | 18.55 (10.51, 24.09) |

| | | | | | |
|---|---|---|---|---|---|
| | | 100% | 23.69 (16.47, 30.11) | -0.44 (-2.73, 1.32) | 23.28 (15.85, 29.98) |
| | 2 | 20% | 2.88 (-0.23, 6.26) | -0.08 (-1.65, 1.55) | 2.75 (-0.26, 6.16) |
| | | 50% | 7.03 (3.03, 11.66) | -0.30 (-1.94, 1.05) | 6.87 (3.00, 11.73) |
| | | 80% | 11.60 (7.15, 18.62) | -0.66 (-2.77, 0.82) | 11.42 (6.89, 18.48) |
| | | 100% | 15.27 (11.27, 21.38) | -0.69 (-3.00, 1.19) | 14.89 (10.81, 20.99) |
| | 3 | 20% | 0.72 (-0.07, 1.45) | -0.03 (-0.49, 0.26) | 0.68 (-0.13, 1.45) |
| | | 50% | 1.71 (0.79, 2.83) | -0.13 (-0.76, 0.40) | 1.65 (0.79, 2.77) |
| | | 80% | 2.87 (1.94, 4.18) | -0.20 (-0.82, 0.43) | 2.80 (1.78, 4.02) |
| | | 100% | 3.76 (2.67, 5.07) | -0.25 (-0.79, 0.46) | 3.59 (2.67, 5.01) |
| | 5 | 20% | 0.03 (-0.10, 0.13) | 0.00 (-0.07, 0.03) | 0.03 (-0.07, 0.13) |
| | | 50% | 0.08 (-0.07, 0.20) | 0.00 (-0.10, 0.10) | 0.07 (-0.07, 0.20) |
| | | 80% | 0.12 (-0.07, 0.33) | 0.00 (-0.10, 0.07) | 0.10 (-0.07, 0.30) |
| | | 100% | 0.13 (-0.07, 0.33) | -0.03 (-0.13, 0.10) | 0.13 (-0.07, 0.33) |
| Deaths averted (thousand) | 1.2 | 20% | 18.68 (-14.14, 58.52) | 14.39 (-19.47, 48.11) | 30.61 (1.69, 71.15) |
| | | 50% | 44.82 (-8.82, 117.86) | 43.09 (1.58, 91.31) | 76.15 (35.78, 146.51) |
| | | 80% | 74.85 (18.23, 148.21) | 65.21 (17.88, 121.49) | 112.83 (53.68, 166.28) |
| | | 100% | 91.66 (20.40, 165.92) | 79.51 (17.71, 143.13) | 136.69 (60.98, 185.92) |
| | 1.5 | 20% | 14.26 (-49.99, 78.46) | 38.59 (-13.91, 86.89) | 46.74 (-1.93, 114.14) |
| | | 50% | 38.24 (-48.24, 121.17) | 91.04 (5.56, 164.45) | 123.80 (39.24, 201.48) |
| | | 80% | 65.85 (-71.70, 162.12) | 157.28 (66.01, 238.88) | 188.03 (89.04, 277.98) |
| | | 100% | 91.43 (-48.31, 178.02) | 190.37 (83.38, 276.20) | 229.81 (124.93, 335.46) |
| | 1.7 | 20% | 12.06 (-44.68, 78.58) | 49.07 (7.54, 104.72) | 57.21 (9.13, 129.86) |
| | | 50% | 39.19 (-62.19, 134.72) | 122.22 (28.96, 197.79) | 146.27 (45.60, 221.34) |
| | | 80% | 57.73 (-43.96, 174.65) | 194.46 (94.44, 297.07) | 226.97 (115.52, 315.25) |
| | | 100% | 68.08 (-40.05, 211.69) | 241.77 (136.21, 349.11) | 273.67 (180.06, 392.08) |

| | | | | | |
|---|---|---|---|---|---|
| | 2 | 20% | 9.04 (-78.52, 74.64) | 63.85 (5.45, 117.52) | 70.77 (4.05, 144.58) |
| | | 50% | 17.11 (-76.91, 129.39) | 155.12 (64.08, 240.41) | 177.71 (83.69, 271.33) |
| | | 80% | 43.22 (-75.05, 166.76) | 247.00 (108.42, 341.85) | 280.87 (158.47, 377.52) |
| | | 100% | 59.20 (-94.86, 173.98) | 316.94 (158.23, 408.09) | 342.26 (209.58, 456.58) |
| | 3 | 20% | 9.96 (-109.69, 76.28) | 100.85 (41.24, 174.82) | 110.70 (35.95, 179.83) |
| | | 50% | 22.20 (-91.08, 123.79) | 256.56 (148.62, 362.85) | 271.85 (156.71, 362.77) |
| | | 80% | 32.03 (-113.81, 158.07) | 411.52 (266.90, 515.39) | 430.13 (277.20, 552.68) |
| | | 100% | 34.60 (-109.69, 182.87) | 507.44 (359.15, 646.19) | 533.39 (384.94, 663.72) |
| | 5 | 20% | 6.94 (-120.85, 116.60) | 153.89 (66.30, 259.38) | 165.92 (56.01, 261.83) |
| | | 50% | 0.92 (-175.10, 154.01) | 388.95 (231.08, 547.49) | 397.71 (246.21, 562.19) |
| | | 80% | 3.40 (-263.34, 205.53) | 625.17 (444.13, 801.75) | 625.01 (447.52, 812.49) |
| | | 100% | -4.43 (-310.29, 245.90) | 761.46 (569.70, 997.59) | 774.24 (586.95, 1002.60) |
| NMB (billion USD) | 1.2 | 20% | 26.57 (-32.77, 103.74) | 19.59 (-35.19, 84.22) | 52.16 (2.62, 122.63) |
| | | 50% | 77.25 (-24.60, 205.73) | 65.99 (-6.92, 151.96) | 131.09 (62.52, 261.32) |
| | | 80% | 124.23 (26.22, 254.08) | 102.74 (13.07, 189.75) | 194.23 (97.62, 293.82) |
| | | 100% | 160.47 (22.57, 288.36) | 121.93 (16.21, 228.25) | 234.46 (107.17, 332.85) |
| | 1.5 | 20% | 15.36 (-90.17, 134.55) | 58.89 (-19.35, 127.21) | 77.31 (-10.22, 194.06) |
| | | 50% | 50.32 (-99.51, 193.46) | 147.49 (-1.33, 276.58) | 199.29 (50.16, 327.12) |
| | | 80% | 83.89 (-140.08, 282.27) | 250.15 (85.13, 389.40) | 307.04 (125.44, 449.21) |
| | | 100% | 118.80 (-99.49, 299.60) | 292.20 (111.03, 447.24) | 376.99 (198.82, 560.47) |
| | 1.7 | 20% | 6.21 (-82.30, 133.89) | 73.95 (5.94, 157.33) | 94.68 (8.54, 196.23) |
| | | 50% | 34.32 (-130.16, 170.20) | 190.44 (32.17, 332.79) | 233.57 (80.45, 379.51) |
| | | 80% | 60.17 (-112.13, 257.44) | 310.34 (109.99, 451.75) | 363.75 (176.38, 499.36) |
| | | 100% | 79.59 (-102.48, 328.03) | 373.38 (196.17, 547.91) | 442.04 (266.32, 610.84) |
| | 2 | 20% | 8.65 (-122.98, 126.50) | 95.27 (3.28, 194.97) | 108.14 (4.57, 246.24) |

| | | | | | |
|---|---|---|---|---|---|
| | 3 | 50% | 19.70 (-169.44, 191.31) | 244.03 (78.62, 396.23) | 287.36 (97.96, 459.36) |
| | | 80% | 53.19 (-184.27, 232.06) | 401.20 (139.63, 559.60) | 441.55 (208.27, 618.77) |
| | | 100% | 66.01 (-194.98, 282.01) | 500.72 (205.19, 687.67) | 546.77 (309.83, 764.58) |
| | 3 | 20% | 5.41 (-172.88, 125.58) | 154.02 (62.97, 284.89) | 174.14 (60.49, 286.14) |
| | | 50% | 2.41 (-179.49, 193.58) | 399.52 (219.88, 578.78) | 426.16 (208.34, 580.13) |
| | | 80% | 11.66 (-239.00, 234.46) | 637.73 (384.92, 820.60) | 675.18 (414.82, 878.32) |
| | | 100% | 3.50 (-272.37, 270.19) | 791.11 (541.89, 982.78) | 826.21 (595.37, 1059.33) |
| | 5 | 20% | -15.94 (-212.90, 142.87) | 229.78 (101.47, 417.79) | 247.30 (92.89, 409.43) |
| | | 50% | -45.26 (-348.55, 188.46) | 586.30 (351.54, 838.32) | 615.21 (366.77, 863.68) |
| | | 80% | -74.83 (-500.47, 261.01) | 956.36 (646.44, 1267.53) | 983.44 (685.66, 1298.68) |
| | | 100% | -104.20 (-612.92, 327.29) | 1183.92 (860.17, 1567.85) | 1209.56 (885.29, 1605.70) |
| Treatment courses used (million) | 1.2 | 20% | 5.75 (4.38, 7.08) | 6.47 (4.78, 7.68) | 5.75 (4.38, 7.15) |
| | | 50% | 12.16 (7.31, 14.89) | 16.49 (12.55, 18.42) | 12.19 (8.86, 14.89) |
| | | 80% | 15.32 (5.93, 20.92) | 26.44 (21.35, 29.42) | 15.44 (6.79, 21.02) |
| | | 100% | 15.78 (4.94, 21.78) | 33.01 (26.66, 36.80) | 15.86 (5.93, 21.91) |
| | 1.5 | 20% | 11.70 (10.31, 12.88) | 11.98 (10.84, 12.88) | 11.71 (10.35, 12.92) |
| | | 50% | 27.64 (25.60, 29.39) | 29.74 (27.51, 31.73) | 27.66 (25.57, 29.36) |
| | | 80% | 41.48 (38.68, 44.88) | 47.56 (44.88, 50.28) | 41.52 (38.78, 45.04) |
| | | 100% | 49.39 (45.17, 53.01) | 59.62 (56.41, 63.26) | 49.54 (45.34, 52.98) |
| | 1.7 | 20% | 13.53 (12.42, 15.09) | 13.82 (12.62, 15.42) | 13.53 (12.42, 15.12) |
| | | 50% | 32.82 (31.04, 34.73) | 34.65 (32.55, 36.70) | 32.88 (30.87, 34.76) |
| | | 80% | 50.94 (47.61, 53.54) | 55.60 (52.09, 58.38) | 50.91 (47.61, 53.90) |
| | | 100% | 61.61 (58.02, 63.95) | 69.47 (65.30, 73.05) | 61.66 (58.22, 64.28) |
| | 2 | 20% | 16.46 (14.73, 17.96) | 16.62 (14.83, 18.09) | 16.44 (14.73, 18.02) |

| | | | | | |
|---|---|---|---|---|---|
| | | 50% | 40.03 (37.96, 42.40) | 41.35 (39.11, 44.35) | 40.02 (37.86, 42.47) |
| | | 80% | 62.80 (58.68, 66.36) | 66.39 (62.57, 70.02) | 62.98 (58.58, 66.19) |
| | | 100% | 77.48 (72.98, 81.94) | 83.16 (78.45, 87.68) | 77.58 (72.98, 81.81) |
| | 3 | 20% | 24.32 (22.31, 26.62) | 24.45 (22.50, 26.72) | 24.30 (22.34, 26.56) |
| | | 50% | 60.10 (57.00, 63.13) | 61.40 (58.45, 64.41) | 60.11 (57.07, 63.16) |
| | | 80% | 94.91 (91.56, 98.71) | 98.32 (94.92, 101.78) | 95.09 (91.70, 98.68) |
| | | 100% | 118.04 (114.50, 121.78) | 122.67 (119.41, 126.75) | 118.17 (114.89, 122.17) |
| | 5 | 20% | 38.39 (35.98, 41.09) | 38.58 (36.08, 41.15) | 38.43 (35.98, 41.19) |
| | | 50% | 95.37 (91.20, 99.27) | 96.23 (92.29, 100.16) | 95.52 (91.50, 99.34) |
| | | 80% | 151.33 (146.42, 156.47) | 154.00 (150.34, 159.50) | 151.56 (146.62, 156.83) |
| | | 100% | 188.88 (184.81, 194.49) | 193.05 (188.47, 198.65) | 189.12 (185.27, 195.02) |
| Hospitalizations reduced (million) | 1.2 | 20% | 0.13 (-0.13, 0.53) | 0.10 (-0.13, 0.40) | 0.26 (0.03, 0.59) |
| | | 50% | 0.40 (-0.07, 0.99) | 0.33 (0.00, 0.76) | 0.66 (0.33, 1.25) |
| | | 80% | 0.63 (0.16, 1.22) | 0.56 (0.10, 0.96) | 0.96 (0.49, 1.42) |
| | | 100% | 0.76 (0.16, 1.35) | 0.68 (0.20, 1.15) | 1.14 (0.56, 1.58) |
| | 1.5 | 20% | 0.10 (-0.40, 0.66) | 0.30 (-0.03, 0.63) | 0.40 (-0.03, 0.92) |
| | | 50% | 0.30 (-0.40, 0.96) | 0.76 (0.07, 1.38) | 1.00 (0.30, 1.58) |
| | | 80% | 0.51 (-0.43, 1.38) | 1.27 (0.49, 1.91) | 1.55 (0.66, 2.14) |
| | | 100% | 0.68 (-0.36, 1.52) | 1.52 (0.66, 2.24) | 1.86 (0.99, 2.67) |
| | 1.7 | 20% | 0.07 (-0.33, 0.66) | 0.40 (0.07, 0.79) | 0.49 (0.07, 0.92) |
| | | 50% | 0.26 (-0.49, 0.86) | 0.99 (0.26, 1.68) | 1.19 (0.49, 1.85) |
| | | 80% | 0.36 (-0.43, 1.32) | 1.58 (0.69, 2.27) | 1.83 (0.89, 2.44) |
| | | 100% | 0.49 (-0.36, 1.61) | 1.94 (1.15, 2.73) | 2.21 (1.38, 2.93) |
| | 2 | 20% | 0.07 (-0.46, 0.59) | 0.49 (0.03, 0.92) | 0.56 (0.10, 1.19) |
| | | 50% | 0.20 (-0.72, 1.02) | 1.22 (0.46, 1.94) | 1.42 (0.59, 2.24) |

| | | | | | |
|---|---|---|---|---|---|
| | | 80% | 0.40 (-0.76, 1.19) | 2.04 (0.86, 2.80) | 2.26 (1.12, 3.00) |
| | | 100% | 0.46 (-0.69, 1.42) | 2.52 (1.19, 3.43) | 2.70 (1.65, 3.72) |
| | 3 | 20% | 0.07 (-0.69, 0.59) | 0.77 (0.40, 1.32) | 0.86 (0.36, 1.38) |
| | | 50% | 0.13 (-0.63, 1.12) | 1.99 (1.15, 2.83) | 2.09 (1.12, 2.83) |
| | | 80% | 0.23 (-0.82, 1.35) | 3.16 (2.08, 4.05) | 3.31 (2.14, 4.22) |
| | | 100% | 0.30 (-0.89, 1.48) | 3.97 (2.90, 4.84) | 4.09 (3.03, 5.11) |
| | 5 | 20% | 0.00 (-0.89, 0.72) | 1.15 (0.59, 2.04) | 1.25 (0.49, 1.94) |
| | | 50% | 0.03 (-1.32, 1.05) | 2.93 (1.88, 3.99) | 3.05 (1.94, 4.25) |
| | | 80% | -0.02 (-1.91, 1.61) | 4.81 (3.26, 6.19) | 4.91 (3.59, 6.36) |
| | | 100% | -0.05 (-2.14, 1.98) | 5.91 (4.48, 7.74) | 6.06 (4.68, 7.78) |

**Appendix Table 5. The optimal choice for Paxlovid treatment under a range of SARS-CoV-2 transmission, testing and isolation scenarios.** We estimated the median (95% CrI) of the number of cases infected averted (millions), number of deaths averted (thousands), number of hospitalizations averted (million), number of courses administered (millions), and net monetary benefit (NMB) in billions of USD, in contrast with baseline, which is scaled to a US population of 328.2 million (13) (**Dataset S1**). Each scenario V1-V3 changes one of the base assumptions, as indicated in the second column. Values in the third column are medians and 95% credible intervals (CrI) in the transmission scenarios ($R_e$ = 1.2 and Treatment rate = 20%) as examples.

| **Scenarios** | | $R_e$ = 1.2<br>Treatment rate = 20% |
|---|---|---|
| | | Incremental net monetary benefits ($ billion), median (95% CrI) |
| Base | Log relationship between infectiousness and viral load | 52.16 (2.62, 122.63) |
| V1 | Log-proportional relationship between infectiousness and viral load[+] | 64.24 (-22.99, 147.37) |
| V2 | Step relationship between infectiousness and viral load[&] | 65.45 (0.71, 141.95) |

| | | |
|---|---|---|
| V3 | Sigmoid relationship between infectiousness and viral load* | 54.75 (-24.03, 125.87) |

+ Infectiousness is proportional log10 of viral load for values above 106, as given by log10(Viral load)-6, and is set to zero otherwise (34).

& Infectiousness is a constant for viral loads above 106, and is set to zero otherwise (34).

* Infectiousness has the sigmoid relationship with viral load following the association between viral load and cell culture isolation success rate (35).

9. Du Z, Pandey A, Bai Y, C. Fitzpatrick M, Chinazzi M, Pastore y Piontti A, et al. Comparative Cost-Effectiveness of SARS-CoV-2 Testing Strategies [Internet].

papers.ssrn.com. 2020 [cited 2020 Nov 14]. Available from: https://papers.ssrn.com/abstract=3714642

20. Tindale, Lauren C and Stockdale, Jessica E and Coombe, Michelle and Garlock,

Emma S and Lau, Wing Yin Venus and Saraswat, Manu and Zhang, Louxin and Chen, Dongxuan and Wallinga, Jacco and Colijn, Caroline. Evidence for transmission of COVID-19 prior to symptom onset. Elife. 2020 Mar 6;9(medrxiv;2020.03.03.20029983v1):e57149.

30. Pouwels KB, Pritchard E, Matthews PC, Stoesser N, Eyre DW, Vihta KD, et al. Effect of Delta variant on viral burden and vaccine effectiveness against new

SARS-CoV-2 infections in the UK. Nat Med. 2021 Dec;27(12):2127–35.
